# Genetic liability to major psychiatric disorders contributes to multi-faceted quality of life outcomes in children and adults

**DOI:** 10.1101/2023.01.17.23284645

**Authors:** Yingjie Shi, Barbara Franke, Nina Roth Mota, Emma Sprooten

## Abstract

**Importance:** Psychiatric disorders can have an immense impact on socioeconomic, physical, and social-psychological facets of life. Psychiatric disorders are also highly heritable. Under a liability threshold model, an important question arises as to what extent genetic liability for psychiatric disorders relates to, and possibly impacts on, different aspects of quality of life in the general population.

**Objective:** To characterize the link between psychiatric genetic liability and diverse aspects of quality of life in childhood and adulthood.

**Design, setting, and participants:** We used data from two multi-site, population-based cohorts, i.e. preadolescent children in the USA enrolled at age 9-10 years from the Adolescent Brain Cognitive Development (ABCD) study (N=4,645) and white British adults between age 40-69 years from the UK Biobank (UKB) study (N=377,664). Due to the current limitations of our genetic methods, only data from unrelated individuals of European descent could be included.

**Main outcomes and measures:** To derive robust measures capturing multiple domains of quality of life in each of the cohorts, we integrated an array of measurements of academic, economic, and physical status, as well as social well-being, in a second-level three-factor confirmatory factor analysis. The genetic liabilities to seven major psychiatric disorders were quantified by a set of polygenic scores (PGSs) derived from the largest genome-wide association studies to date, independent of the target cohorts, of major depressive disorder (MDD, N=142k-173k), anxiety disorders (ANX, N=22k-144k), attention-deficit/hyperactivity disorder (ADHD, N=226k), autism spectrum disorder (ASD, N=55k), schizophrenia (SCZ, N=130k), bipolar disorder (BIP, N=353k-414k), and cannabis use disorder (CUD, N=384k). Using general linear models we assessed associations between PGSs and the estimated latent factors, controlling for age, sex, site, genotyping batch, plate, and genetic ancestry.

**Results:** In each cohort, three latent factors indexing distinct but correlated quality of life domains, (1) educational performance and cognition (Edu, in ABCD) / social economic status (SES, in UKB), (2) physical health (Hea), (3) adverse social experience (Adv, in ABCD) / social well-being (Soc, in UKB), were estimated with excellent model fit indices. In addition, a general factor was derived that captured the covariances between the three latent factors (QoL). In the ABCD cohort, ADHD-PGS was significantly associated with Edu (β = -0.13, t = -8.29, p = 1.53e-16), Adv (β = -0.09, t = -5.79, p = 7.81e-09), and general QoL (β = -0.14, t = -8.74, p = 3.37e-18) factors. In the UKB cohort, all examined disorder PGSs were significantly associated with the general QoL latent factor and at least one first-order subdomain, with ADHD-PGS (β = -0.06 ∼ -0.10, t = -29.1 ∼ -52.5, p < 5.91e-186) and MDD-PGS (β = -0.04 ∼ -0.07, t = -23.8 ∼ -36.3, p < 3.63e-125) showing the largest effects.

**Conclusions and relevance:** The present study reveals an inverse relationship between psychiatric genetic liabilities and multiple quality of life metrics, with ADHD-associated genetic risk being the main contributor in both children and adults, and MDD additionally showing effects in adults. All effect sizes observed were small, as expected. Understanding potential real-world outcomes of quantitative measures of disorder-related genetic risks in the general population can provide a scientific foundation for societal intervention and policy-making processes, with profound implications for promoting a flourishing society.

## Introduction

The impact of psychiatric disorders extends beyond the domain of mental well-being, and involves a broad range of aspects of life spanning educational, occupational, physical, social, and psychological outcomes. Epidemiological studies have linked psychiatric disorders with reduced overall quality of life, as well as specific domains of functioning^1,2^. Even though many countries have made quality of life their policy aim, there is no unified way of conceptualizing and measuring this construct. A widely accepted quality of life model acknowledges both the aspects of quantifiable standard of living complying with societal expectations, as well as subjective evaluations of well-being^3^.

The boom of large-scale genome-wide association studies (GWASs) has enabled the identification of common genetic variations contributing to psychiatric disorders such as attention-deficit/hyperactivity disorder (ADHD), autism spectrum disorder (ASD), major depressive disorder (MDD), anxiety disorders (ANX), schizophrenia (SCZ), bipolar disorder (BIP), and cannabis use disorder (CUD). Built on the resultant increasingly accurate genome-wide statistics, individuals’ genetic susceptibility to disorders can be quantified by polygenic scores (PGSs), which have proven useful in risk stratification of common complex diseases^4^. PGSs can also provide genetic risk proxies in cohorts without symptom/trait measures and enable the estimation of different disorders’ contribution to variables of interest.

Here, we aimed to assess how genetic susceptibility for seven major psychiatric disorders indexed by PGSs relates to diverse life-relevant outcomes. We took advantage of two large population cohorts, the Adolescent Brain Cognitive Development (ABCD) study and the UK Biobank study, to study different phases of the lifespan (preadolescent children and middle-aged adults). We derived multi-faceted quality of life constructs capturing general and specific domains of human functioning and experiences at these two life stages.

## Methods

Our study sample consisted of 4,645 non-Hispanic White preadolescent children (47% females, age 9.92±0.62 years) recruited across the United States of America as part of the ABCD study cohort (request 11315, data release 4.0) and 377,664 white British, unrelated adults (54% females, age 56.95±7.94 years) as a subset of the population-based UK Biobank study cohort (application 23668, data release 3.0). Written informed consent was obtained for all participants involved in the UK Biobank study and both parents and children (verbal consent) in ABCD study. Full details of genetic data (pre-)processing, factor model estimation, and statistical analyses are described in **Supplementary Information**.

Seven major psychiatric disorders, which have well-powered GWAS results, with varied ages of onset and symptom presentation, were chosen as the bases for computing PGSs. For each target cohort, we curated the largest and most recent GWAS results, without overlap with the target samples, for ADHD^5^, ASD^6^, MDD^7^, ANX^8,9^, SCZ^10^, BIP^11^, and CUD^12^ (sample sizes shown in **Supplementary Table S1**. PRS-CS^13^ was employed as the primary polygenic scoring method and results were validated across two other methods (i.e., C+T and PRS-PCA).

To extract latent factors representative of key aspects of quality of life at different stages of life, we conducted a confirmatory factor analysis (CFA) in each cohort. For the ABCD study cohort, we aggregated measures in areas of 1) educational performance and cognition (Edu): child- and guardian-reported school grades, cognition composite score; 2) physical health (Hea): doctor visits, disease conditions, emergency room visits, overnight hospital stays; and 3) adverse social experience (Adv): experiences of peer victimization and cyberbullying. For the UK Biobank cohort, indicators spanned the domains of 1) social economic status (SES): household income, educational qualifications; 2) physical health (Hea): self-rated health, long-standing illness and disability, diagnosis of serious medical conditions; and 3) social well-being (Soc): frequency of confiding, loneliness, and excessive worry of embarrassment. Both models contain the above three first-level factors and one second-level QoL factor. Model performance was evaluated based on a variety of fit indices and quality of life latent factors were estimated via an empirical Bayes method using the lavaan package^14^. More details of the measurements included in the analyses are available in **Supplementary Tables S2 and S3**.

Genotyping batch and plate, study site, sex, age, and genetic ancestry were controlled for in the regression models, where the final model fit R^2^ was derived after subtracting the R^2^ of the null model including these covariates. Bonferroni-corrected p values (for 7 PGSs * 4 latent factors * 3 PGS methods = 84 tests) < .05 were considered significant.

## Results

Descriptive statistics of all indicators included in the CFA models are presented in **Supplementary Figure S1**. In the ABCD cohort, the defined second-order model had an excellent model fit (CFI = 0.989, RMSEA = 0.021, SRMR = 0.030, TLI = 0.984) (**Figure 1**, upper panel). A similarly structured second-order model was estimated in the UK Biobank cohort (CFI = 0.971, RMSEA = 0.046, SRMR = 0.043, TLI = 0.952) (**Figure 1**, lower panel). All model parameters are presented in **Supplementary Tables S4 and S5**. In a subset of the UK Biobank sample in which self-rated satisfaction measures were available, the three first-level latent factors estimated from the model structure were significantly associated with satisfaction in their corresponding life domain (**Supplementary Table S6**).

**Figure 1.**
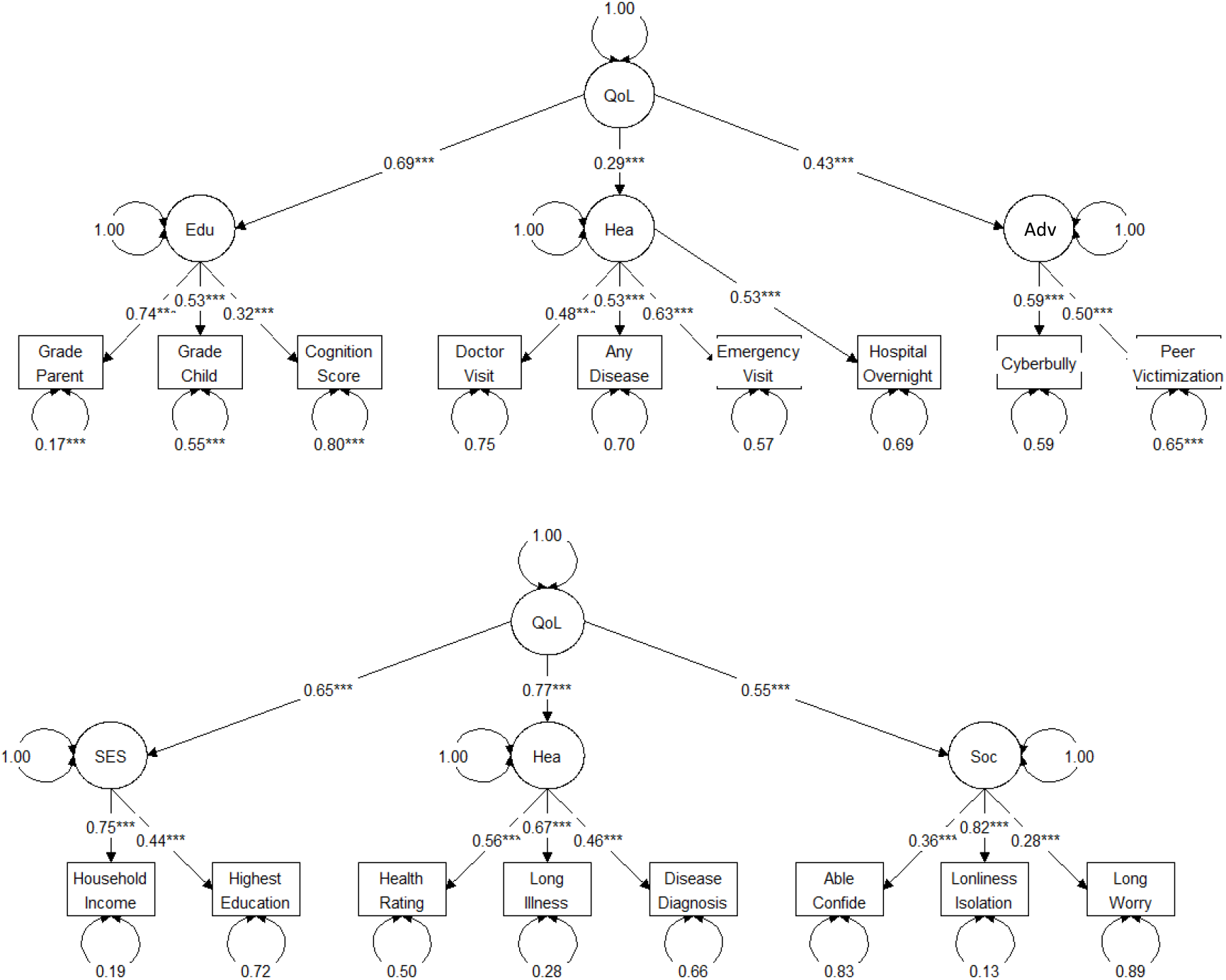
Primary CFA model structures for ABCD (upper) and UK Biobank (lower). Variables have been sign-flipped so that higher scores correspond to higher levels of quality of life. Upper panel: Edu - educational performance and cognition; Hea - physical health; Adv - adverse social experience. Lower panel: SES - social economic status; Hea - physical health; Soc - social well-being.

In the ABCD cohort, PGS for ADHD significantly explained 1.67%, 0.84%, and 1.85% of Edu (β = -0.133, SE = 0.016, t = -8.29, p =1.53e-16), Adv (β = -0.094, SE = 0.016, t = -5.79, p = 7.81e-09), and general QoL factors (β = -0.140, SE = 0.016, t = -8.74, p = 3.37e-18), respectively; PGS for other disorders were not significantly associated with any of the latent factors (**Figure 2**, upper panel, **Supplementary Table S7**). In the UK Biobank cohort, PGSs based on all seven psychiatric disorders were associated with the general QoL latent factor and at least one first-order subdomain (**Figure 2**, lower panel, **Supplementary Table S7**). Among them, ADHD-PGS showed the largest effect and explained the most variance in the general QoL factor (β = -0.096, SE = 0.002, t = -52.5, p < 2.23e-308, R^2^ = 0.009), SES (β = - 0.081, SE = 0.002, t = -46.5, p < 2.23e-308, R^2^ = 0.007), and Hea (β = -0.083, SE = 0.002, t = -43.74, p < 2.23e-308, R^2^ = 0.007); most variances in Soc was explained by the MDD-PGS (β = -0.060, SE = 0.002, t = -31.23, p = 9.96e-214, R^2^ = 0.004).

**Figure 2.**
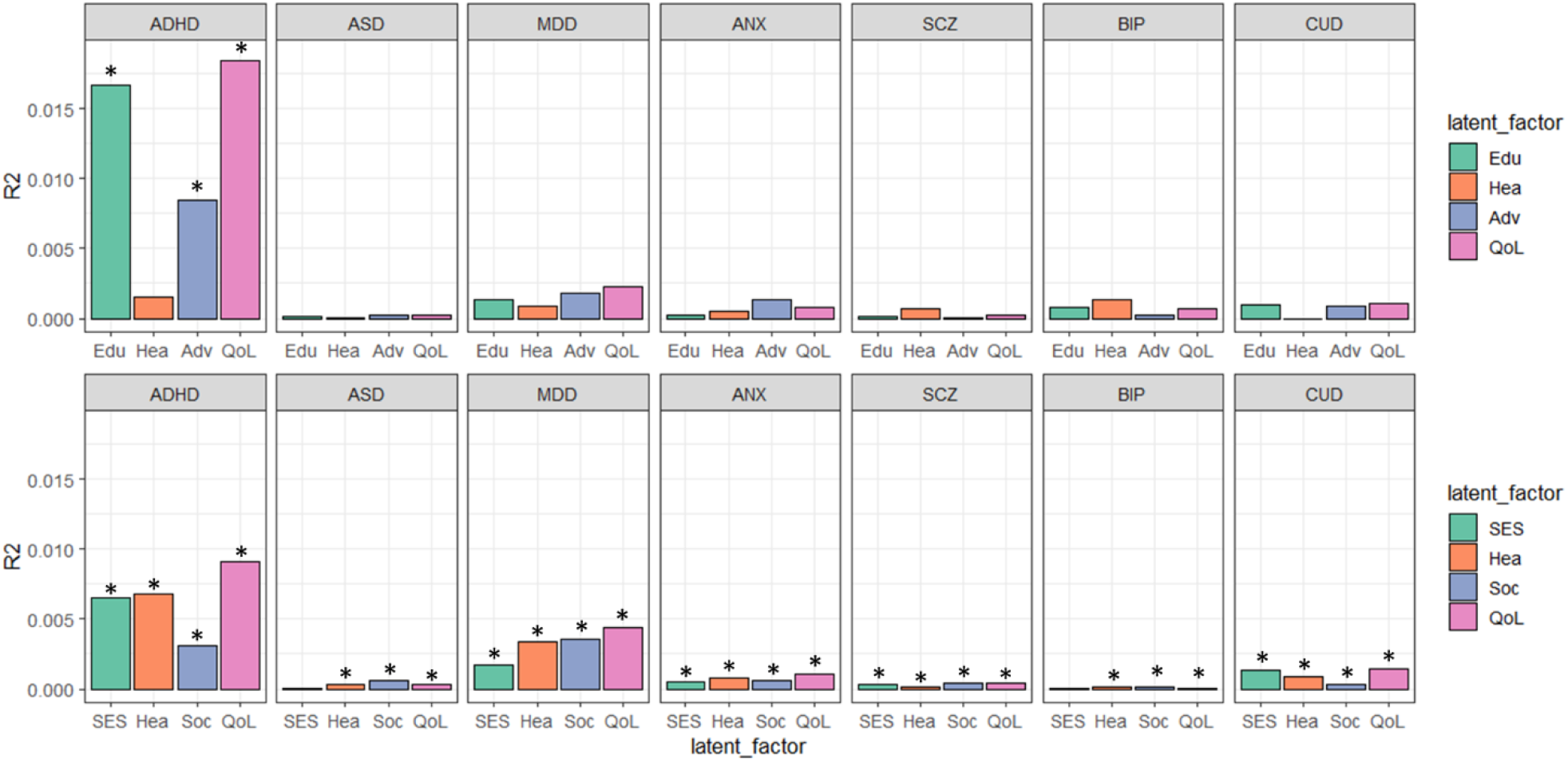
Variance explained by polygenic scores derived from different major psychiatric disorders in quality of life latent factors for 3,909 children from the ABCD study cohort (upper) and 269,293 adults from the UK Biobank study cohort (lower). Upper panel: Edu - educational performance and cognition; Hea - physical health; Adv - adverse social experience; QoL – overall quality of life; Lower panel: SES - social economic status; Hea - physical health; Soc - social well-being; QoL – overall quality of life.

## Discussion

Here, we set out to evaluate the effects of psychiatric genetic liabilities on quality of life in childhood and adulthood. We mapped PGSs encompassing a broad range of psychiatric disorders to diverse, age-specific aspects of quality of life, concerning academic, socio-economic, physical, and social well-being. We found higher ADHD genetic liability to be associated with worse general quality of life, educational performance, and social well-being in early life. In the adult sample, the effect was observed across all seven psychiatric disorders examined, with ADHD-PGS and MDD-PGS showing the largest effects.

We combined quality of life indices across a range of domains and scales, self-perceived and objectively quantified. Empirically derived quality of life constructs that are sensitive to genetic variation pave the way toward personalized treatment goals relevant to individuals’ functioning and adaptations in their own lives and in society, which is complementary to symptom reduction as the current primary indicator of treatment efficacy.

The negative relationships we identified between genetic liability to major psychiatric disorders and quality of life related outcomes were robust, despite their effect sizes being small. The common variants captured by the polygenic scores reflect a fraction of the total heritability of these disorders and can thus only explain a small portion of variance in their primary phenotypes (i.e., disorder status)^7–12^. Nevertheless, the genetic liability to ADHD was associated with both childhood and adulthood outcomes, suggesting that genetic liability to ADHD may be most sensitive to the potential negative impact on quality of life among the psychiatric disorders tested at this moment, and this effect cannot be fully explained by power (**Supplementary Information**). Joint efforts in increasing the diversity and sample sizes of GWASs, as well as exploiting data for rare and copy number variants are essential to provide a more complete individual genetic risk profile for psychiatric disorders.

### Limitations

Prior research^15^ suggested a ‘healthy volunteer selection bias’ in the UK Biobank cohort, where the sample was enriched in wealthier and healthier individuals. This may limit the generalizability of the current results, especially for disorders such as SCZ and ASD, where the debilitating genetic effect might be more pronounced in the samples at the higher end of the liability spectrum. Moreover, this study only provided snapshots of childhood (9-10 years) and part of adulthood. Longitudinal data are helpful to further elucidate how these genetic risks manifest along the trajectory of human development and aging.

## Conclusions

Combining newly available GWAS results with genotyped and richly phenotyped cohorts of children and adults, our results present an inverse relationship between psychiatric genetic liability and multiple aspects of quality of life. Polygenic scores provided a means to evaluate the contributions of genetic liability for different psychiatric disorders to different aspects of life in the general population. The established quality of life constructs could help to identify relevant biological and environmental factors to the mechanisms underlying well-being.

## Supporting information

Supplementary Methods, Results, and Figures 1-5

## Data Availability

Data from the ABCD Study is available at the NIMH Data Archive (NDA). The UK Biobank resource is open to all bona fide researchers under request. Genome-wide association summary statistics are publicly available from psychiatric genomics consortium.

https://www.ukbiobank.ac.uk/

https://nda.nih.gov/

https://pgc.unc.edu/

## Acknowledgements

We thank Prof. Frank Dudbridge for the helpful discussions regarding the power analyses.

