## Supplementary Methods, Results, and Figures 1-5 for "Genetic liability to major psychiatric disorders contributes to multi-faceted quality of life outcomes in children and adults"

**Supplementary Results**

**Supplementary Figures 1-5**

**Supplementary References**

Corresponding email:

**Supplementary Methods**

**Genotyping, Quality Control, and Imputation of the target datasets**

Detailed descriptions of the QC and imputation steps in both cohorts have been provided in previous publications^1,2^. Briefly, UK Biobank participants were genotyped using either the Affymetrix UK BiLEVE Axiom array or the UK Biobank Axiom array^3,4^. A set of stringent quality control (QC) procedures was performed, followed by further imputation using the Haplotype Reference Consortium, UK10K and 1000 Genomes phase 3 reference panels, which resulted in a dataset of 93,095,623 variants.

We further applied sample and variant filtering prior to polygenic score analyses based on the following protocol. We removed individuals with sex mismatch, sample missingness larger than 0.05, heterozygosity outside of 3 standard deviations, and those of non-European descent based on self-report and principal components analysis (PCA) of the genotypes. To account for sample relatedness, we identified related individual pairs with KING kinship coefficient higher than 0.0884 (first- and second-degree relatives) and created family clusters. A greedy algorithm (<https://rdrr.io/cran/ukbtools/man/ukb_gen_samples_to_remove.html>) was applied to trim the connections until only one individual was left within each cluster. Single nucleotide polymorphisms (SNPs) were excluded if they had a Hardy-Weinberg equilibrium (HWE) test P value < 1e−6, genotype missing rate > 0.05, minor allele frequency (MAF) < 0.005, or imputation quality of INFO < 0.8, resulting a final list of 10,203,392 SNPs.

The ABCD cohort was genotyped using the Affymetrix National Institute on Drug Abuse SmokeScreen Array^5^. Standard pre-imputation QC was conducted following the recommendations of the Ricopili pipeline^6^, after which the resulted variants were imputed to the Trans-Omics for Precision Medicine (TOPMed) panel. The 307,467,480 SNPs were further filtered according to the following criteria: imputation R^2^ > 0.8, MAF > 0.005, HWE p > 1e-6, genotype missing rate < 0.05, yielding 10,537,977 variants. A subset of SNPs with MAF > 0.05 and linkage disequilibrium-pruned with 500kb window size and 0.2 r^2^ was included to calculate genetic principal components (PCs). We identified unrelated participants, by removing all but one participant per family, and non-Hispanic white participants, based on guardians’ reports and results from PCA.

**Quality of life outcomes and confirmatory factor analysis**

Given the great phenotyping depth of both cohorts, more than one well-fitting model structures with sets of reasonable indicators were available. Here, for each cohort, we provided one theoretically sound and statistically best-fit model as the primary model used in the main analyses, as described in the main text and presented in **Figure 1**, and we also provided the model parameters of a less well-fitting alternative model, presented in **Figure S3**. **Supplementary Tables S2** and **S3** show a full list of indicators included in the primary or alternative model. The correlation matrix of indicators included in both the primary and alternative models was presented in **Supplementary Figure S2**. To ease the interpretation, some variables were sign-flipped so that larger values always correspond to higher quality of life levels.

In the ABCD cohort, we fitted the model with nine observed variables indicating three latent factors – educational performance and cognition (Edu), physical health (Hea), and adverse peer experience (Adv). Observed variables are, for Edu, school grades in the past year reported by parents (sag_grade_type) and youths (sag_grades_last_yr), cognition total composite score (abcd_tbss01:nihtbx_totalcomp_uncorrected); for Hea, ever seen doctors before the past year (excluding regular check-ups) (abcd_mx01:medhx_1b), ever seen doctor for any (severe) diseases (derived from abcd_mx01), emergency room visits before the past year (derived from abcd_mx01), ever been in the hospital overnight or longer (abcd_mx01:medhx_8a); for Adv, ever been cyberbullied (abcd_cb01:cybb_phenx_harm), experienced victimization from peers (derived from abcd_peq01). Individuals who reported ‘do not know’, ‘not applicable’, or ‘refuse to answer’ to the abovementioned items were excluded from the analyses. A final list of 3,909 participants with phenotypic data available was included in downstream analyses.

In the UK Biobank cohort, we included three latent factors – social economic status (SES), health (Hea), and social wellbeing (Soc) – which were indicated by eight binary and ordinal variables: average total household income before tax (UKB index 738) and educational qualifications (6138) for the first factor; overall health rating (2178), long-standing illness, disability or infirmity (2188), and whether diagnosed with any serious medical conditions (derived by aggregating item 6150, 6152, 2443, 2453, and 2473) for the second factor; the frequency of being able to confide with someone (2110), whether often feel loneliness or in isolation (2020), and the tendency to worry too long after embarrassment (1930) for the third factor. For educational qualifications, we converted each individual’s highest qualification to an International Standard Classification of Education (ISCED) category and removed other qualifications that were not included in such classification scheme. Individuals who reported ‘do not know’ or ‘prefer not to answer’ to the abovementioned items were excluded from the analyses.

The CFA model was implemented in lavaan package^7^ using the method of weighted least squares mean and variance adjusted (WLSMV), which used diagonally weighted least squares to estimate model parameters, but the full weight matrix to compute robust standard errors. The variances of all latent variables (i.e., ‘factor scores’) in the models were fixed to unity, and their estimated values were computed using empirical bayes method (EBM). The metrics of Comparative Fit Index (CFI), Root Mean Square Error of Approximation (RMSEA) and 90% confidence intervals, Standardized Root Mean Square Residual (SRMR), together with Tucker-Lewis Index (TLI) were employed to assess the model fit. CFI values above 0.95, RMSEA values below 0.06, SRMR values below 0.08, and CFI values above 0.95 were considered as evidence for good model fit^8^.

**Polygenic scores calculation and statistical analysis**

Polygenic scores for disorders of interest were computed using a linkage disequilibrium r^2^ threshold of 0.1 and a sliding window of 250 kb. A set of eight P-value thresholds (P_T_ = 1e-6, 1e-4, 0.001, 0.01, 0.05, 0.1, 0.5, 1) were specified and the scores were derived and compared using three approaches: 1) classical clumping and thresholding with the p value that yields the best fit (C+T)^9,10^; 2) a principal component approach (PRS-PCA)^11^; and 3) a Bayesian-based continuous shrinkage method (PRS-CS-auto)^12^, as reported in main text.

We held out around one-eighth of the UK Biobank sample as an independent tuning set (N = 37,766) in the C+T approach, where the p threshold that gave the best model fit was selected and evaluated in the test set (N = 269,293). To keep the sample size consistent, the association tests of the scores derived from PRS-PCA and PRS-CS methods were also conducted in the same test set. Given the relatively smaller sample size in ABCD study (N = 3,909 with complete PGSs and estimated factor scores), we did not further split this cohort and optimized the threshold using the full sample in the C+T approach. For the PRS-PCA approach, the first principal component of polygenic scores (PRS-PC1) derived from *a priori* specified thresholds reweighted the variants for maximum variation. In the PRS-CS approach, a global shrinkage parameter was automatically learned from GWAS summary statistics and imposed on the SNP effect sizes. If not otherwise specified, the results in the main text were based on PRS-CS, and regression results for all three approaches were presented in **Supplementary Figures S4** and **S5**.

Simple linear regression was performed between each polygenic score (as predictor) and QoL latent factor (as outcome). Sex, age in years, batch, site, the first ten ancestry informative genotype PCs for the UK Biobank cohort and batch, site, plate, sex, age at baseline in months, and the first ten ancestry informative genotype PCs for the ABCD cohort were included as covariates. The model fit R^2^ was defined as the variances explained by the null model containing only covariates subtracted from the full model comprising one polygenic score and all covariates. Multiple regression models with all 7 PGSs were also constructed to assess the overall variance explained by the different PGSs altogether. Bonferroni correction was applied accounting for the number of polygenic scores and outcome variables tested (7 PGSs * 4 latent factors * 3 PGS methods = 84 tests), with the corrected p value < 0.05 being considered significant.

Given the high genetic correlation between ADHD and intelligence, as a sensitivity analysis, to rule out the possibility that the effect of ADHD-PGS was confounded by intelligence, we constructed the PGS for IQ based on Savage et al. (2018)^13^ and added the polygenic scores for intelligence as the covariate.

**GWAS of QoL factors and genetic correlations with psychiatric disorders**

We further conducted genome-wide association (GWA) analyses on the four estimated latent factors in the UK Biobank cohort using PLINK 2^14,15^, with an assumed additive genetic model. The association analyses were adjusted for sex, age in years, batch, site, and the first ten genotype PCs. We derived SNP heritability estimates of each latent factor and computed their genetic correlation with the 7 psychiatric disorders using linkage disequilibrium score regression (LDSC) v1.0.1^16^.

**PGS power analyses**

To give an indication of the statistical power of the PGS analyses, we conducted a power calculation using the ‘avengeme’ package^17^ in both cohorts and provided the estimates in **Supplementary Tables S8** and **S9** at different genetic correlations (r_g_ = 0.2, 0.4, 0.6, 0.8, 1.0). The calculation was based on the classical C+T method, which has been shown to be a good approximation^18^ to the joint modelling of correlated variants and can be used as a rough lower bound for the power estimates using the PRS-CS approach. Since the SNP heritability of the latent factors from the ABCD cohort was unknown, we estimated this parameter based on previous study of similar traits. The estimates of SNP heritability of the disorders and population prevalence were obtained from the original GWAS publications. We assumed 5% of SNPs to have an effect on the base trait, and the power estimates were maximized across the specified p thresholds.

**Supplementary Results**

**GWAS of QoL factors and genetic correlations with psychiatric disorders**

GWA analysis in UK Biobank (N = 377,664) revealed SNP heritability for four latent factors (SES: h^2^_SNP_ = 0.080, Hea: h^2^_SNP_ = 0.067, Soc: h^2^_SNP_ = 0.045, QoL: h^2^_SNP_ = 0.082), which were in line with estimates for the individual indicators^19^. All seven PGSs together explained 10.8% of SNP heritability of SES, 13.1% of Hea, 12.7% of Soc and 14.8% of general QoL. Variance explained proportional to SNP heritability by single PGSs are presented in **Supplementary Table S10**. LD score regression analyses revealed significant genetic correlations between all seven disorders and at least one QoL subdomain (**Supplementary Table S11**). In particular, ADHD, MDD and ANX are highly (negatively) genetically correlated with all the QoL factors.

**Robustness of results and sensitivity analyses**

Overall, the results were consistent across the polygenic scoring method of C+T, PRS-PCA and PRS-CS-auto (**Supplementary Figures S4** and **S5**). In the UK Biobank cohort, in which the sample size is sufficiently large for such PGS analysis (as suggested by the power analysis), we see a gradual increase in variances explained by the PGS methods of C+T, PRS-PCA and PRS-CS-auto, accordingly. This pattern is most prominent in PGSs for ADHD, MDD, and CUD, for which the effects were the largest. In the ABCD study cohort, where the sample size is relatively small, PRS-CS-auto boosted the R^2^ of ADHD-PGS to a large extent, with regard to Edu and QoL outcome, but did not improve the R^2^ in other associations too much.

Previous research has established a moderate to high genetic correlation between ADHD and cognition-related outcomes (e.g., intelligence^13^, *r*_g_ = −0.36, *p* = 4.58 × 10^−23^; educational attainment, r_g_ = -0.515, SE = 0.025, p < 10^-6^). As a sensitivity analysis, we examined the associations in both cohorts between ADHD-PGS and QoL outcomes after adding PGS for intelligence into the model. The effects of ADHD-PGS on all QoL factors remained significant after covarying for IQ-PGS. **Supplementary Table S12** shows the unique and shared variance explained by the two variables.

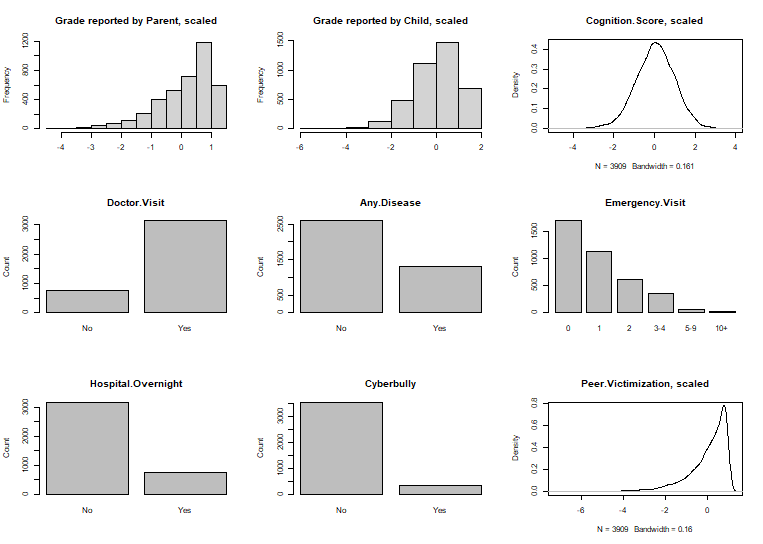

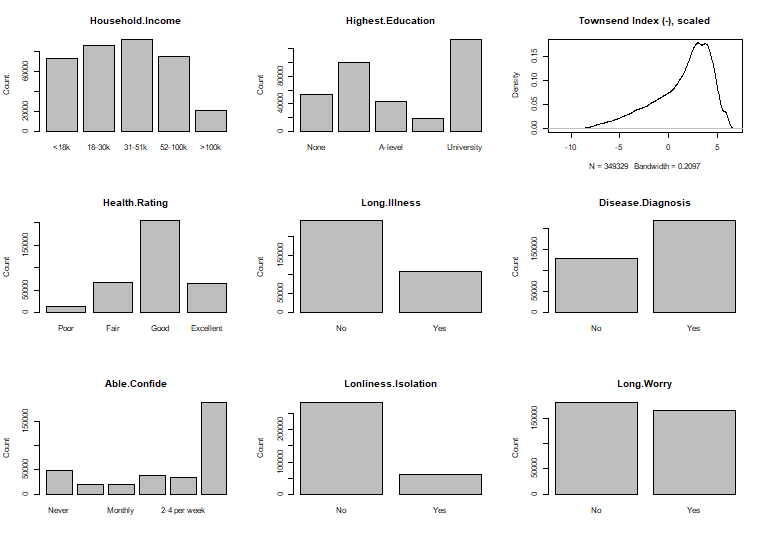

**Figure S1.** Distributions of all indicators (included in the primary and/or alternative CFA models) in ABCD (left) and UK Biobank (right) cohorts.

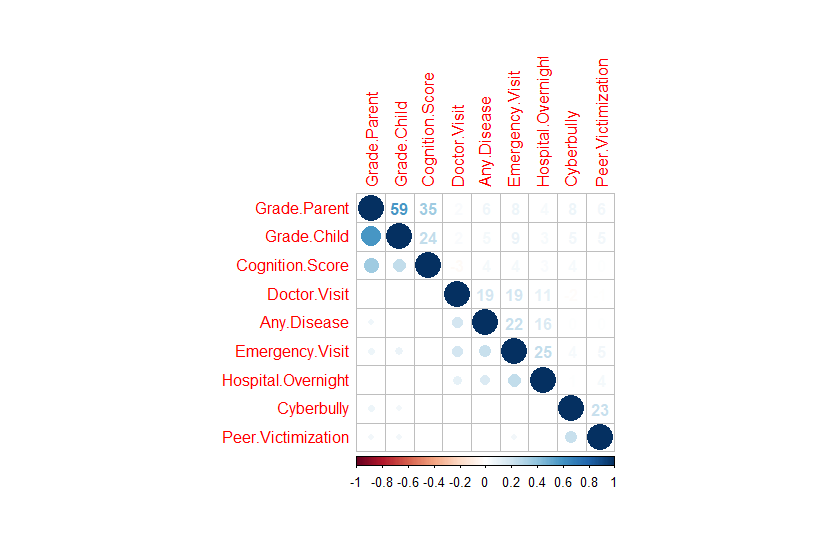

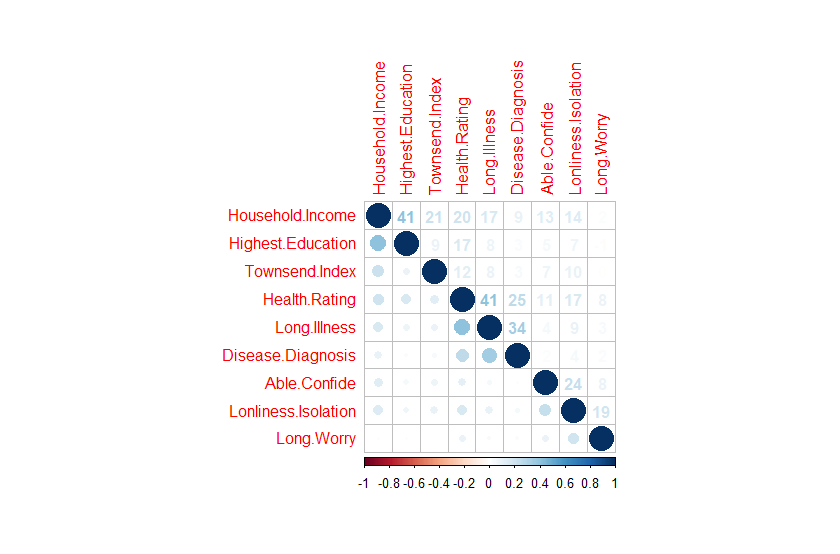

**Figure S2**. Correlation matrices of all indicators (included in the primary and/or alternative CFA models) in ABCD (left) and UK Biobank (right) cohorts. Spearman’s correlation coefficients are shown as percentages on the upper diagonal.

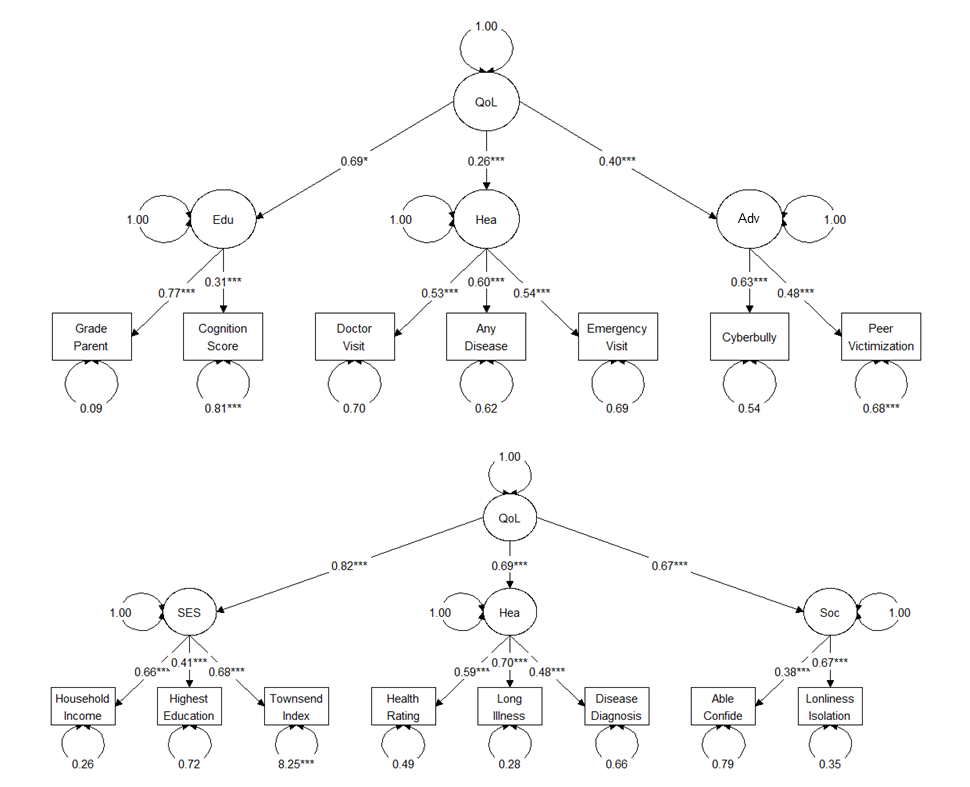

**Figure S3.** Alternative CFA model structures for ABCD (upper) and UK Biobank (lower). ABCD: CFI = 0.977, RMSEA = 0.031, SRMR = 0.032, TLI = 0.956. UKB: CFI = 0.969, RMSEA = 0.049, SRMR = 0.048, TLI = 0.948.

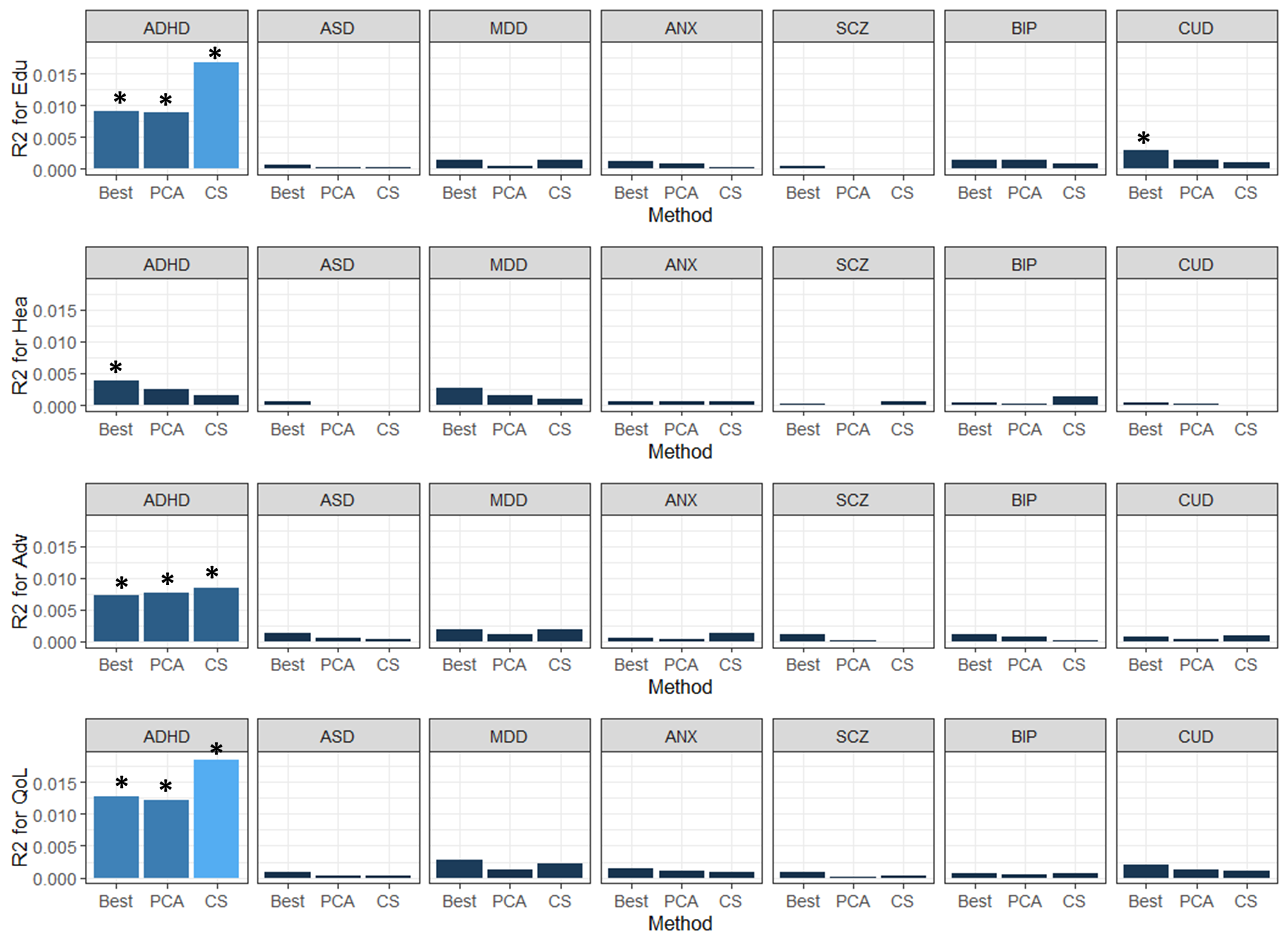
**Figure S4.** Variance explained by polygenic scores derived through three methods in quality of life latent factors for ABCD cohort. Best: clumping and thresholding with the p value that yields the best fit; PCA: the first principal component of polygenic scores across eight pre-specified thresholds; CS: continuous shrinkage of effect sizes with the global shrinkage parameter automatically learnt from the data. Asterisk indicates significance after multiple test corrections.

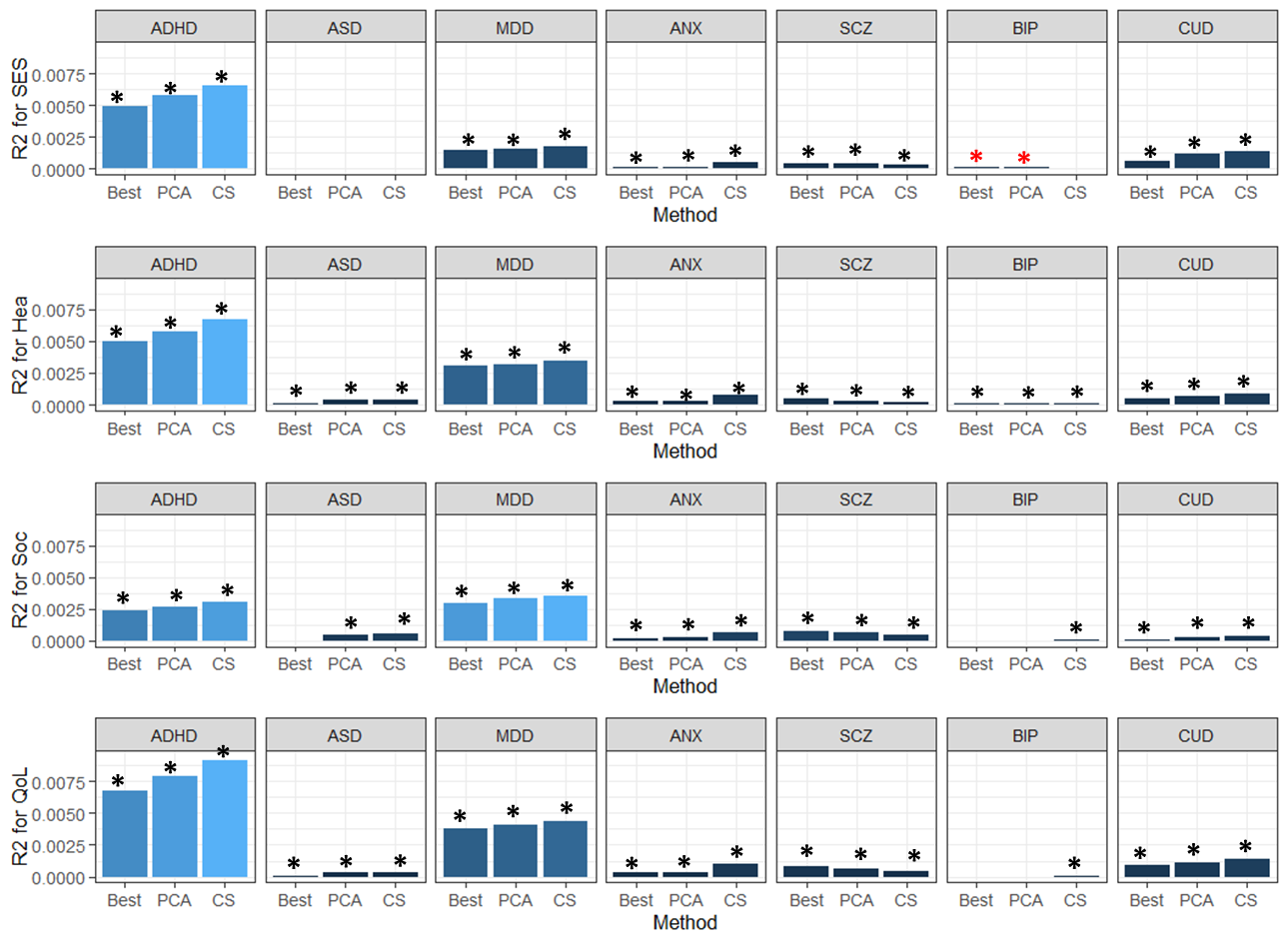

**Figure S5.** Variance explained by polygenic scores derived from three approaches in QoL latent factors for UK Biobank. Best: clumping and thresholding with the p value that yields the best fit; PCA: the first principal component of polygenic scores across eight pre-specified thresholds; CS: continuous shrinkage of effect sizes with the global shrinkage parameter automatically learnt from the data. Asterisk indicates significance after multiple test corrections. Red asterisk indicates significant effect in the opposite direction, i.e., higher PGS, higher quality of life outcome.
